# Network based statistics shows that rem sleep behavior disorder and visual hallucinations increase functional connectivity in early dementia with Lewy bodies

**DOI:** 10.1101/2025.02.26.25322839

**Authors:** Laura Carini, Sara Sommariva, Francesco Famà, Laura Giorgetti, Pietro Mattioli, Beatrice Orso, Raffaele Mancini, Matteo Pardini, Michele Piana, Dario Arnaldi

**Author notes:** Correspondence to: Sara Sommariva.

## Abstract

Dementia with Lewy Bodies (DLB) is a heterogeneous disease characterized by four core clinical features, namely visual hallucinations, REM sleep behaviour disorder (RBD), cognitive fluctuations and parkinsonism. In this paper, we perform for the first time a comprehensive study on the relationship between all these features and functional connectivity derived from high-density electroencephalographic data. To quantify functional connectivity, we used two different connectivity metrics, each one averaged on individual theta and alpha frequency bands. The latter were defined by determining the individual theta-to-alpha transition frequency through a previously validated package called transfreq. The study was performed using a cohort comprising 33 subjects affected by early stage DLB (mean age 82 ± 6 years, males/females: 23/10) and 21 healthy controls (mean age 71 ± 9 years, males/female: 10/11).

After showing that overall DLB determines a shift toward lower frequencies of posterior dominant rhythm and theta-alpha transition, we used Network Based Statistic (NBS) to explore differential connectivity networks between subgroups of DLB patients with different core features. We consistently found that the presence of both visual hallucinations and RBD is associated with increased connectivity in early DLB patients mostly in the left hemisphere, while cognitive fluctuations and parkinsonism appear to have a non-significant impact on functional connectivity metrics. These findings may represent an initial compensatory mechanism in response to underlying neurodegeneration.

## Introduction

Dementia with Lewy Bodies (DLB), which represents the second most common cause of neurodegenerative dementia after Alzheimer’s Disease (AD), is an age-related form of dementia presenting the formation of Lewy bodies in both brain-stem nuclei, and in paralimbic and neocortical regions^1^. Indeed, DLB patients are characterized by specific core features reflecting the involvement of both cortical and sub-cortical structures, such as cognitive fluctuations, visual hallucinations, parkinsonism^1^ and REM sleep behaviour disorder (RBD)^1^. Some of these features, particularly RBD, can emerge years before the onset of dementia^2,3^. Indeed, RBD is now considered a prodromal stage of alpha-synucleinopathies, including DLB^4,5^.

The early cortical dysfunction characterizing DLB patients can be efficiently assessed using EEG, especially when advanced functional connectivity metrics are applied to high-density EEG (hdEEG) data. Overall, network connectivity is typically reduced in DLB compared to healthy controls, with some possible increase specifically in the frontal-temporal network^6^. However, characterising differences in connectivity between DLB patients with different clinical features as well as between DLB and other neurodegenerative diseases is still an open problem^7,8^. Most literature data focused on the relationship between functional connectivity metrics and cognitive functions and/or visual hallucinations^6^. Recent neurophysiological studies have provided initial empirical evidence for the possibility of modelling functional networks related to visual hallucinations in DLB^9,10^. Nevertheless, other core features of DLB may also be associated with cortical dysfunctions that manifest as altered functional connectivity metrics.

For example, modifications in alpha phase synchronisation and delta amplitude correlation may represent an electrophysiological signature of cortical compensatory mechanisms in iRBD patients. This suggests that large-scale functional networks could serve as early biomarkers for alpha-synucleinopathies^11^. Moreover, EEG-based machine learning models have successfully identified iRBD patients at high risk of short-term phenoconversion and tracked their longitudinal trajectories. Indeed, patients exhibiting more altered connectivity metrics are more likely developing DLB^12^. However, there is a lack of comprehensive literature assessing the relationship between functional brain connectivity dysfunctions and all core features of DLB.

In EEG signal analysis, standard frequency bands, such as delta ([1,4]) Hz), theta ([4,8] Hz, alpha ([8,13] Hz) and beta ([13, 30] Hz) bands, are commonly used to investigate brain activity in both healthy and pathological conditions. When dealing with healthy subjects, these bands typically succeed in capturing the corresponding brain rhythms although they may vary considerably according to factors such as age, memory performance, brain volume and task demands^13,14^. Instead, these standard frequency ranges may be inaccurate applied to subjects affected by neurodegenerative disorders. For instance, the Dominant Posterior Alpha Brain Rhythm, which typically falls within the standard alpha band for healthy subjects, may shift to lower frequencies in subjects with DLB^15^, risking misclassification within the standard theta band. To address this issue, Klimesh et al.^16^ introduced a data–driven approach for computing the individual theta-to-alpha transition frequency (TF), which represents the frequency that discerns the end of the theta band from the beginning of the alpha band. Different approaches have been developed to improve Klimesh’s method by leveraging different physiological criteria^16–18^.

To fill these gaps of the currently available literature on DLB, in this study we utilize individual frequency bands to explore the relationship between each core clinical features of DLB (visual hallucinations, cognitive fluctuations, RBD, and parkinsonism) and abnormalities in functional brain connectivity. Through this work we confirm the capability of high-density electroencephalography (hdEEG) to provide new insights into these associations.

## Results

### Demographic data and clinical heterogeneity

Our cohort included 33 patients affected by early DLB (82 ± 6 years), and 21 HC (71 ± 9 years) whose main demographic data and clinical scores are summarised in Table 1. All DLB patients had a positive presynaptic dopaminergic imaging as an indicative biomarker and/or a positive brain glucose imaging as a supportive biomarker^1^. Two patients were using low dose benzodiazepines at the time of EEG recording. Due to the limited sample size of our cohort we decided to also keep these two subjects in our analysis. However, we reran all the analysis after excluding those two patients and the results confirmed the trends discussed throughout the paper.

**Table 1.** Demographic data and clinical scores for DLB and HC.

|  | DLB (33) | HC (21) |
| --- | --- | --- |
| Age | 82 ± 6 years | 71 ± 9 years |
| Male/Female | 23/10 | 10/11 |
| MMSE | 22 ± 6 | - |
| MDS-UPDRS-III | 18 ± 13 | - |
| Education | 9 ± 5 | 13 ± 4 |
| Dominant Hand (R/L) | 28/5 | - |

Continuous variables are shown as mean ± standard deviation.
*DLB* Dementia with Lewy Body, *HC* Health Control, *MMSE* MiniMental State Examination score, *MDS-UPDRS-III* Movement Disorders Society-sponsored revision of the Unified Parkinson's Disease Rating Scale

The UpSet Plot^19^ in Figure 1 describes the distribution of the core clinical features within the DLB sample. In detail, cognitive fluctuations were present in 9 subjects, RBD in 19 subjects, visual hallucinations in 22 subjects, and parkinsonism was found in 28 subjects. One subject did not present any of the considered core clinical features. Although this patient had a polysomnography-confirmed RBD before the diagnosis of DLB, at the time of EEG recording the RBD was not clinically evident. Thus, this patient had the RBD core clinical feature for DLB diagnosis purposes, but we considered this patient as without any core feature for the subsequent analysis. However, we also ran all the analyses by excluding this patient, and our results were confirmed.

**Figure 1.**
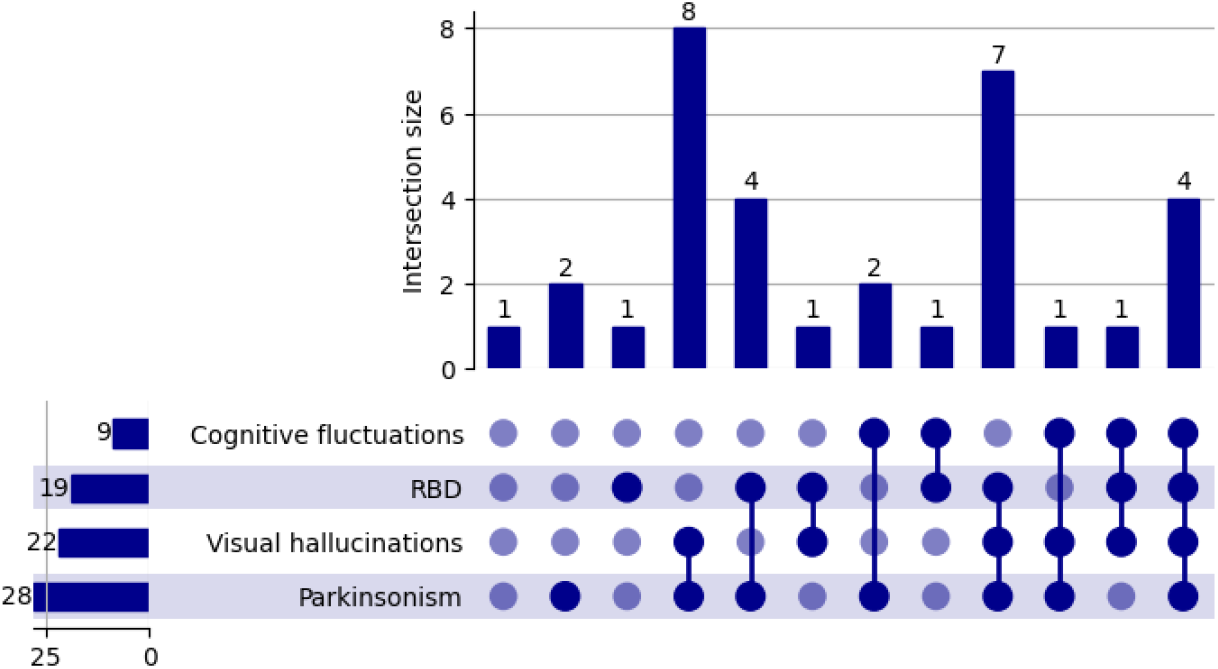
Distribution of the core clinical features and of their intersection within the cohort of DLB patients. Bar chart on the left expresses the number of subjects presenting a clinical feature; the bar chart on the top shows the size of each intersection set. Each row corresponds to a possible intersection: the filled-in cells show which set is part of an intersection.

### Individual rhythm analysis confirms the shift to lower values of the transition frequency for DLB subjects

For each DLB patient and each healthy control we used a modified version of the 2-dimensional k-means algorithm, introduced by Vallarino and colleagues^18^, to group the EEG channels based on the corresponding averaged spectral density in the theta and alpha bands. For each subject we defined two groups of channels, denoted as *G*_*θ*_ and *G*_*α*_, showing a high value of the averaged spectral density in the theta and in the alpha band, respectively. Topographical maps in Figure 2 compare the results obtained by carrying out this clustering procedure using the standard frequency bands with the results obtained by using the individual frequency bands defined through transfreq as described in the Material and Methods section. For DLB subjects, posterior-occipital channels, expected to more accurately reflect the Dominant Posterior Alpha Rhythm, were clustered in the *G*_*α*_ group when using the individual band definition and in the *G*_*θ*_group when applying the standard band definition. This confirms the shift to lower frequencies of the Dominant Posterior Alpha Rhythm. In contrast, in the case of healthy controls, both the standard and individual analyses successfully clustered the posterior-occipital channels in the *G*_*α*_ group.

**Figure 2.**
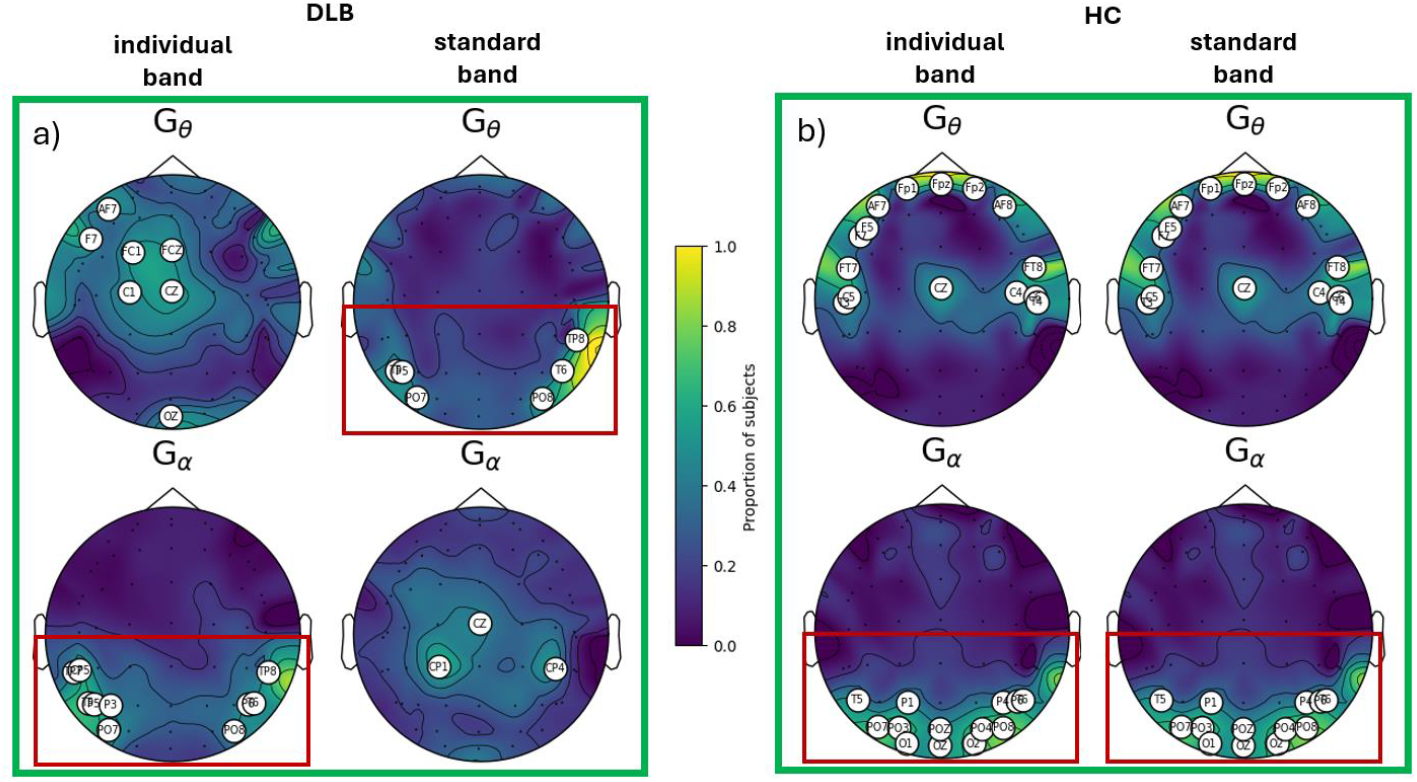
Topographical maps visualising for each EEG channel the number of subjects for which that channel is grouped into *G*_*θ*_, as it shows a high content in the theta band (upper row), or into *G*_*α*_, a s it shows a high content in the alpha band (lower row). Panel a) shows the results for DLB patients, while panel b) concerns healthy controls. In each panel, the theta and alpha bands are defined using the individual analysis (left column) and the standard definition (right column). In each map we highlighted the channels for which the number of subjects is greater than 40%.

These results underline the importance of using individual frequency bands to study brain rhythms, particularly in subjects affected by neurodegenerative diseases. For this reason, in the remaining of the paper we investigated connectivity by averaging the values of weighted phase lag index (WPLI) and the absolute value of imaginary part of coherency (|IMCOH|) within the individual theta and alpha bands.

### Clinical heterogeneity in DLB patients reflects in hdEEG functional dysconnectivity when compared to HC

To infer differences in hdEEG functional connectivity induced by DLB, we used NBS to test the null hypothesis of equal connectivity between the cohort of DLB patients and the HC. Furthermore, for each of the four core clinical features assessed here, we divided the DLB patients in two groups: those presenting the feature and those who did not. As above, we used NBS for group-level comparison of the connectivity in each subgroup of DLB patients and the HC.

Table 2 shows the tests that returned a statistically significant (p < 0.05) differential network for at least one metric between WPLI and |IMCOH|. We observe that the whole cohort of DLB patients showed a reduced connectivity than HC when using the averaged value of WPLI in the individual alpha band. Figure 3(a) shows that such a differential network, consisting of 188 connections between 51 nodes, spreads across the whole brain suggesting a general impairment of functional connectivity in DLB patients. However, this result was not confirmed when using |IMCOH| as a connectivity metric. Also, DLB patients without visual hallucinations (DLB(Visual hallucinations-)) showed a lower value of WPLI in the individual alpha band than HC. The corresponding differential network, shown in Figure 3(b), spreads across the whole brain as in the previous comparison involving the whole DLB cohort. On the other hand, DLB patients with RBD (DLB(RBD+)) and with visual hallucinations (DLB(Visual hallucinations+)) separately showed increased values of connectivity in the individual alpha-band while DLB patients with cognitive fluctuation (DLB(Cognitive fluctuations+)) showed an enhanced connectivity in the individual theta-band. Notably, the only result confirmed by both the considered connectivity metrics is the one concerning DLB patients with RBD. The corresponding differential networks are shown in Figure 3(c) for WPLI and in Figure 3(d) for |IMCOH|. Both networks showed connectivity from the right posterior regions to the contralateral fronto-temporal areas. However, WPLI seems to be more conservative than |IMCOH| as a lower number of nodes were involved in the corresponding network.

**Table 2.** Network based statistics for comparing DLB patients with HC.

|  |  | WPLI | \|IMCOH\| |
| --- | --- | --- | --- |
|  | DLB < HC | p = 0.021 | n.s. |
| Individual αband | DLB(Visual hallucinations-) < HC | p = 0.020 | n.s. |
|  | DLB(RBD+) > HC | p = 0.049 | p = 0.015 |
|  | DLB(Visual hallucinations+) > HC | n.s. | p = 0.036 |
| Individual $\theta$ band | DLB(Cognitive fluctuations+) > HC | p = 0.017 | n.s. |

Results of NBS applied on WPLI and |IMCOH| connectivity matrices within individual frequency bands, using the alternative hypothesis of a greater (>) or smaller (<) connectivity in groups of DLB patients than in HC. The presence or absence of a specific core clinical feature is denoted with + or -, respectively. P-values and “n.s.” are used to mark tests that did or did not return a statistically significant ( $p < 0.05$ ) differential network. Only tests returning a statistically significant differential network for at least one connectivity metric are shown.

**Figure 3.**
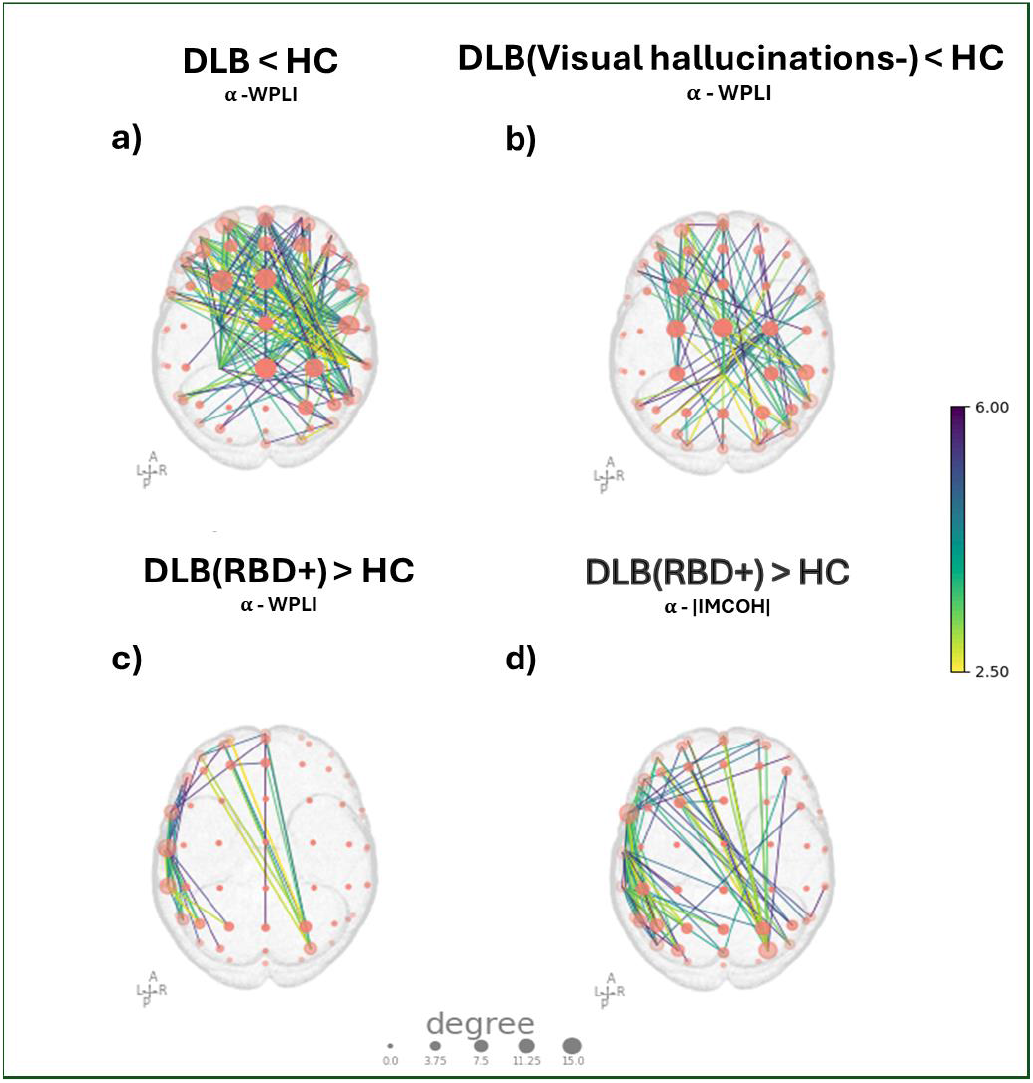
Statistically significant differential networks provided by NBS when comparing connectivity matrices of DLB patients and of HC. a) Results of testing for a diminished value of WPLI within the individual alpha band in the whole cohort of DLB patients than in HC. b) Results of testing for a diminished value of WPLI in the individual alpha band of DLB patients without visual hallucinations (DLB(Visual hallucinations-)) than in HC. c)-d) Results of testing for increased value of WPLI and |IMCOH|, respectively, in the individual alpha band from DLB patients with RBD (DLB(RBD+)) than in HC. In each panel, the edge colours represent the t-values provided by the first step of NBS while each node size is proportional to its degree.

For the sake of conciseness, the differential networks for DLB patients with visual hallucination and with cognitive fluctuations, that are statistically significant for only one connectivity metric, are shown in Supplementary Figures 1 and 2, respectively. The presence/absence of parkinsonism did not significantly impact the functional connectivity metrics.

### RBD and visual hallucinations increase functional connectivity in DLB patients

We then investigated the effect of the four core clinical features (RBD, visual hallucinations, cognitive fluctuations, and parkinsonism) on the functional connectivity metrics of DLB patients. We used NBS to test the null hypothesis of equal connectivity between the same two subgroups considered in the previous section i.e., we performed one test for each core feature using the average value of WPLI and |IMCOH| in the individual theta and alpha bands. NBS did not yield any statistically significant differential network when testing for greater connectivity in patients without the symptom. Instead, Table 3 summarises the results obtained when the alternative hypothesis was that connectivity is greater in patients with the symptom compared to those without. Patients with RBD presented a greater connectivity in the individual alpha-band than patients without RBD, while enhanced theta-band connectivity was found in patients with visual hallucinations as compared with those without visual hallucinations. Both results were confirmed using either WPLI or |IMCOH|. The connected graphs differentiating DLB patients with RBD from those without RBD are shown in Figure 4(a) and consisted in 71 connections between 30 nodes when WPLI is used, while |IMCOH| showed 81 connections between 39 nodes. Furthermore, in both cases the node degree fell in a range between 0 and 15, with higher values for fronto-temporal and posterior nodes. The violin plots in Figure 4(b) quantitatively compare the connectivity for the two groups: the value of both WPLI and |IMCOH| was, on average, higher for the DLB patients with RBD (WPLI mean = 0.39 ± 0.05; |IMCOH| mean = 0.09 ± 0.02) than for patients without RBD (WPLI mean = 0.27 ± 0.03; |IMCOH| mean = 0.04 ± 0.01). Figure 5(a) shows the connected graphs differentiating DLB patients with visual hallucinations from the other. When WPLI was used, the graphs consisted in 108 connections between 47 nodes, while when |IMCOH| is used, the graph involved 32 connections between 25 nodes. Furthermore, the violin plots in Figure 5(b) confirmed that the value of WPLI and |IMCOH| is, on average, higher for patients with visual hallucinations (WPLI mean = 0.31 ± 0.02; |IMCOH| mean = 0.06 ± 0.01) than for patients without hallucinations (WPLI mean = 0.24 ± 0.01; |IMCOH| mean = 0.03 ± 0.006).

**Table 3.** Network based statistics for comparing groups of DLB patients presenting different clinical features.

| | Individual $\theta$ band | | Individual $\alpha$ band | |
| --- | --- | --- | --- | --- |
|  | WPLI | \|IMCOH\| | WPLI | \|IMCOH\| |
| DLB(RBD+) > DLB(RBD-) | n.s. | n.s. | p = 0.042 | p = 0.020 |
| DLB(Visual hallucinations+) > DLB(Visual hallucinations-) | p = 0.017 | p = 0.026 | n.s. | n.s. |

Results of NBS applied on WPLI and |IMCOH| connectivity matrices computed for the individual frequency bands. Null hypothesis for each core clinical feature: equal connectivity in patients presenting the feature and those who did not. Alternative hypothesis: enhanced connectivity in patients presenting the feature. p-values and “n.s.” mark tests that did or did not return a statistically significant ( $p < 0.05$ ) differential network. Only tests returning a statistically significant differential network for at least one connectivity metric are shown.

**Figure 4.**
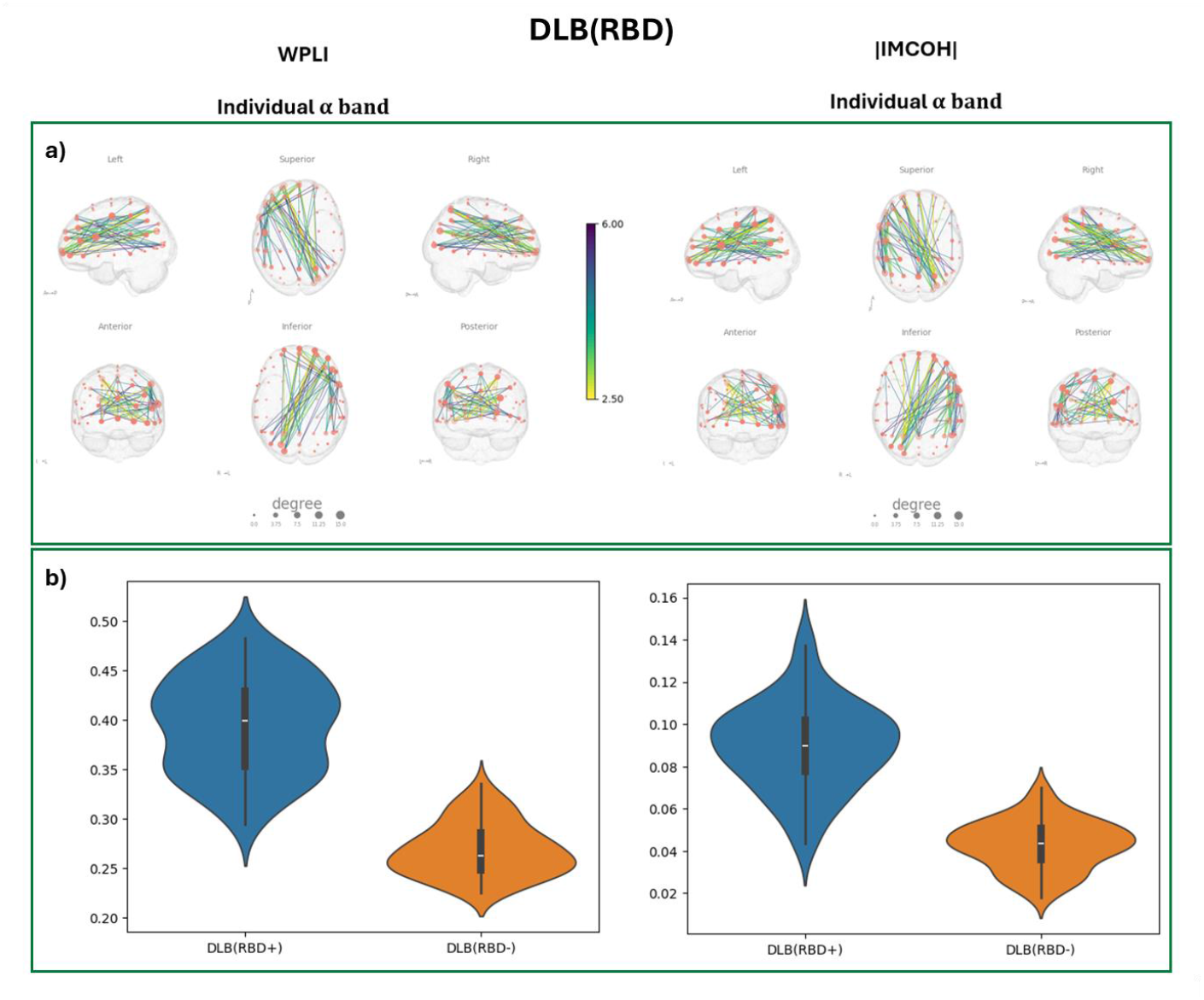
Differences in functional connectivity in the individual alpha band between DLB patients with and without RBD. a) Outcome of NBS when testing the alternative hypothesis of a greater connectivity in DLB patients with RBD (DLB(RBD+)) than in DLB patients without RBD (DLB(RBD-)). The colour of the edges represents the t-values obtained from the first univariate test in NBS while the size of each node is proportional to its degree. b) Distribution of the connectivity metrics’ values across subgroups at the edges of the graphs in a). In each panel the first column refers to the results obtained using WPLI while the second column concerns |IMCOH|.

**Figure 5.**
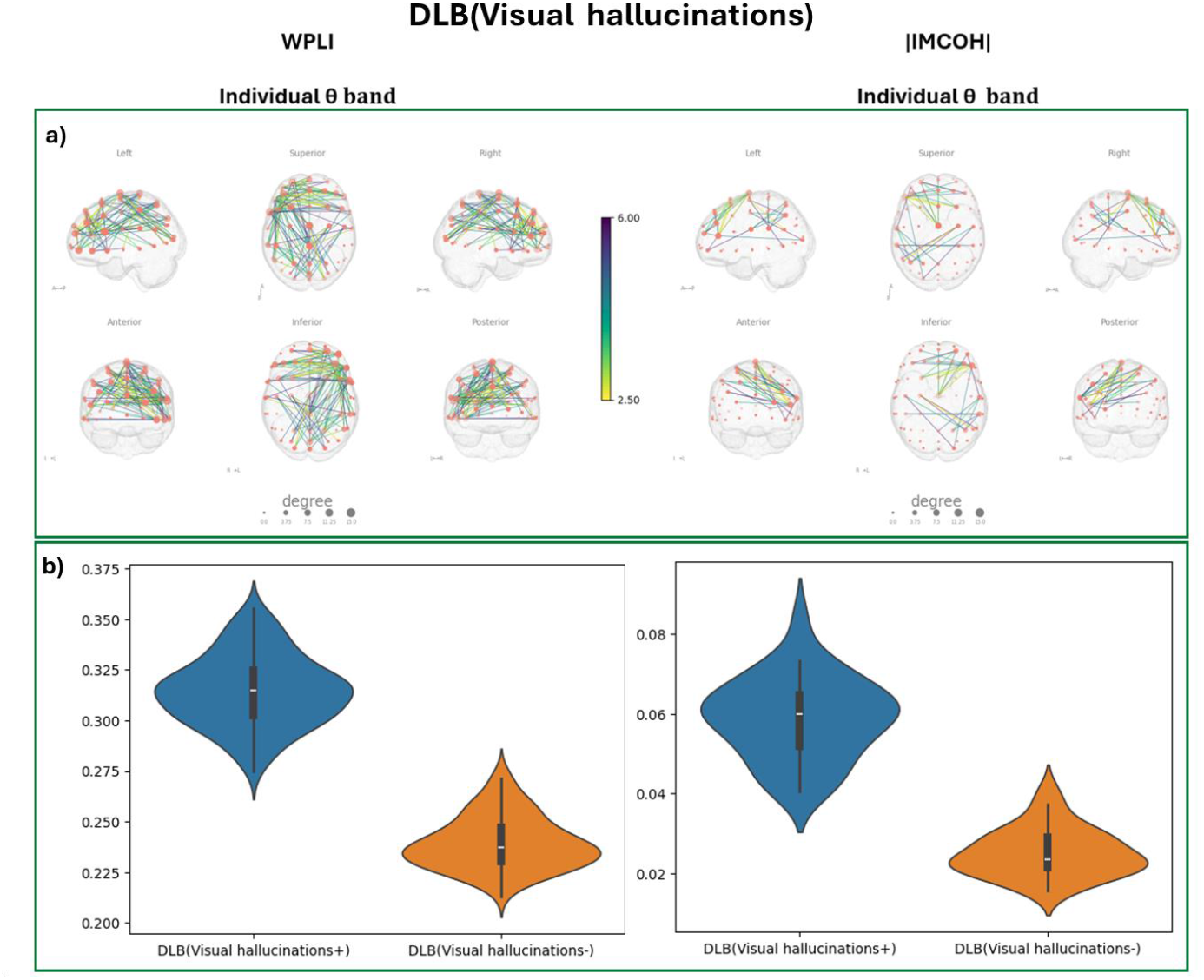
Differences in functional connectivity in the individual theta band between DLB patients with and without visual hallucinations. a) Outcome of NBS when testing the alternative hypothesis of a greater connectivity in DLB patients affected by visual hallucinations (DLB(Visual hallucinations+)) than in the other DLB patients (DLB(Visual hallucinations-)). The colour of the edges represents the t-values obtained from the first univariate test in NBS while the size of each node is proportional to its degree. b) Distribution of the connectivity metrics’s values across subgroups at the edges of the graphs in a). In each panel the first column refers to results obtained using WPLI while the second column cncerns |IMCOH|.

The presence/absence of parkinsonism and cognitive fluctuations did not significantly affect functional connectivity metrics.

## Discussion

For the first time, in this study, we comprehensively investigated the relationship between functional connectivity metrics, assessed through network based statistics in hdEEG data, and all the core clinical features characterizing DLB patients, namely visual hallucinations, RBD, cognitive fluctuations and parkinsonism. We consistently demonstrated, using various metrics and approaches, that the presence of both visual hallucinations and RBD is associated with increased connectivity in early DLB patients, mostly in the left hemisphere. Conversely, DLB patients showed overall reduced functional connectivity compared to healthy controls. Cognitive fluctuations and parkinsonism seem to have a non-significant impact on functional connectivity metrics.

Several studies have investigated functional connectivity in DLB patients, mainly using functional MRI and EEG, as well as DTI and FDG-PET^6,20^. All studies agree that there is a global reduction in connectivity metrics in DLB patients compared to healthy subjects^6,20^, and we confirm this finding also in our sample. However, literature data show conflicting results regarding the direction of connectivity when different hypothesis-driven approaches are employed, as well as in patients at different stages of the disease^6,20^.

A meaningful drawback of most previous literature is the reliance on standard frequency bands. However, it is known that neurodegenerative diseases affecting the central nervous system, especially DLB, cause a shift toward lower frequencies of both the posterior dominant rhythm and the theta-to-alpha transition frequency^21^. We confirm this finding also in our sample of DLB patients, and therefore we applied the network based statistic to individual frequency bands, instead of standard ones, likely increasing the reliability of our findings. In fact, the use of standard frequency bands in DLB patients is critical, as it poses the risk of combining subjects with non-homogeneous individual EEG characteristics. For instance, the posterior dominant rhythm may fall within either the standard alpha or theta range, potentially leading to inconsistent results due to the specific characteristics of the study cohort. To note, in the present cohort, the transition frequency consistently fell within the standard theta frequency band, thus in line with existing literature^22^. This finding suggests that using the standard alpha and theta frequency bands may not have captured the individual characteristics of the evaluated subjects.

Furthermore, most literature focuses on specific, a priori defined networks, such as the default mode network. While this approach is valuable when a strong a priori hypothesis is made, it may not be the best choice for wider and more innovative aims. In the present study, we chose a whole brain approach to investigate the unexplored relationship between all four DLB core features and functional connectivity metrics.

Additionally, it is important to emphasize the importance of using individual frequency bands rather than standard ones, also for topographic reasons. Indeed, as can be seen in the Figure 2, in DLB patients, the standard alpha frequency band is not expressed in the typical parieto-occipital regions, where instead standard theta is highly represented. This discrepancy has relevant implications when interpreting both network-based and region-based analyses applied on standard frequency bands, as typically shown in the current literature.

Interestingly, in the present study we found that the presence of both visual hallucinations and RBD was associated with increased functional connectivity in the individual theta and alpha bands, respectively. Notably, those findings were confirmed by applying two different functional connectivity measures, namely the weighted phase lag index (WPLI) and the absolute value of imaginary part of coherency (|IMCOH|). The main difference between these two connectivity metrics is that while WPLI is a measure of phase synchronization, the value of |IMCOH| depends on both phase and amplitude correlation^23^. From an electrophysiological point of view, phase synchronisation reflects the communication between coherently oscillating spatially distant neuronal groups^24^. Instead, amplitude correlation in EEG signals has been related to the concurrent activation of different neuronal populations triggered by a sensory stimulus^11^.

It has been postulated that brain networks work in a balanced state to maintain efficient information integration as well as their functional state^25,26^. Any deviation from this balanced state, in either direction, implies an irregularity. Thus, the increased connectivity associated with visual hallucinations and RBD may represent an initial compensatory mechanism in response to the underlying neurodegeneration process. To note, all enrolled DLB patients were in an early disease stage, thus likely reflecting an initial neurodegeneration phase. Supporting this hypothesis, increased functional connectivity has also been found in patients with isolated/idiopathic RBD^11^, thus indicating an even earlier alpha-synucleinopathy stage. Conversely, several studies have found reduced connectivity metrics associated with visual hallucinations. However, these studies evaluated more heterogeneous cohorts that included not only DLB patients, but also PD patients with dementia (PDD), who are likely in a different, more advanced stage of the disease. Additionally, these studies used standard frequency bands instead of individual ones^9,27^. Finally, the presence of increased connectivity as a compensatory mechanism in the early neurodegeneration stages has been suggested also in the field of Alzheimer’s disease^28,29^. Another possible physiopathological interpretation of the increased connectivity is that the repeated presence of abnormal clinical manifestations, such as visual hallucinations and RBD, determine stressful effects on brain networks that, in response, react with increased activity, phenomenon that has been speculated to occur in psychiatric disorders characterized by hallucinations^30^.

In the present study, both core features (visual hallucinations and RBD) were associated with increased connectivity across a broad network involving brain regions included in the fronto-parietal network, as well as additional temporal and central brain areas. Interestingly, this increase primarily affected the left (dominant) hemisphere. This is in agreement with existing literature that demonstrates widespread brain network dysfunctions when a whole brain approach is applied, rather than relying on predefined networks^31–33^, particularly when considering individual rather than standard frequency bands^34^. Furthermore, in agreement with our findings, a previous, large, multicentric brain [^18^F]-FDG-PET study showed that the presence of visual hallucinations and RBD was associated with a relative increase in brain glucose metabolism, especially in the temporal lobe^35^.

Notably, most previous literature on functional connectivity studies has focused on visual hallucinations, while we found that the presence of RBD significantly affects brain functional connectivity as well. This is in line with existing literature showing that presence of RBD from the prodromal to overt alpha-synucleinopathy continuum is associated with an early cortical involvement^11,12,36,37^. Conversely, neither cognitive fluctuations nor parkinsonism significantly affected the functional connectivity metrics in our sample.

This study has some limitations such as a relatively small sample size, as well as its single-modality nature, which means that except for hdEEG no other biomarkers of brain function, such as [^18^F]FDG-PET or fMRI, were used.

Despite these limitations, in this study we carefully characterized our sample based on the biological definition of the disease, by including patients with an abnormal presynaptic dopaminergic imaging. Moreover, all patients underwent overnight polysomnography which is considered the gold standard for diagnosis of RBD. Furthermore, we used individual frequency bands, computed using a validated semi-automatic tool^18^, likely enhancing the consistency and reliability of our results. Finally, this study is the first to investigate the relationship between functional connectivity and all four core clinical features of DLB.

## Materials and methods

### Participants and experimental protocol

The original data set included patients affected by DLB who underwent hdEEG. Participant recruitment took place at the IRCCS Ospedale Policlinico San Martino in Genova, Italy. Diagnoses were performed by following international guidelines^1^, and all patients had at least one positive indicative biomarker, specifically presynaptic dopaminergic imaging and/or brain glucose imaging. Furthermore, Mini-mental State Examination (MMSE) and the movement disorder society Unified Parkinson’s Disease Rating Scale motor examination (MDS-UPDRS-III) were used to assess global cognition and motor signs, respectively. The presence of visual hallucinations and cognitive fluctuations was inferred from clinical evaluation conducted by an experienced clinician. RBD was diagnosed using overnight polysomnography, serving as an additional indicative biomarker in these cases. In order to have all binary features in the statistical analysis, a patient was considered to have parkinsonism based on clinical evaluation and when the value of the MDS-UPDRS-III was equal or greater than 6.

For comparison, subjects without neurological or psychiatric disorders (healthy controls, HC) were selected from the lab internal database matched for age to the DLB patients.

All participants signed an informed consent form in compliance with the Helsinki Declaration of 1975. The study was approved by the local institutional board.

### EEG data acquisition and preprocessing

Recording sessions were performed at the IRCCS Ospedale Policlinico San Martino using a standard high-density EEG cap with 64 electrodes during an eyes-closed resting state task. For both DLB patients and HC, recordings lasted around 15-20 minutes and took place in a dimly lit and silent room where participants sit, awake, at rest with closed eyes. From each EEG recording, a time-series of 5-6 mins was extracted by an expert EEG technician by visually inspecting the EEG recordings and selecting possibly discontinuous segments without evident artifacts. Recordings were digitised at 512 Hz and the EEG cap presented a reference electrode between FZ and AFZ.

Preprocessing of the EEG data was performed using the MNE-Python analysis package^38^. First, a Hamming-windowed finite impulse response (FIR) filter was applied to each EEG recording with a range from 1 to 40 Hz. Then, time series were visually inspected and bad channels were manually removed and then interpolated (number of removed channels: 4 ± 3). Data were re-referenced using average reference^39^ and independent component analysis (ICA)^40^ was performed to remove noise and artefacts such as eye blinks, heartbeats, and muscular contractions (number of removed components: 9 ± 3).

### Individual and standard frequency bands

In this study we applied Transfreq^18^ (source code available at https://github.com/elisabettavallarino/transfreq), a Python package that allows for the computation of the theta-to-alpha transition frequency (TF) from resting state data by clustering the spectral profiles associated with EEG channels based on their content in the alpha and theta bands. We chose this method because it requires only one EEG time-series recorded at rest, while all other available approaches also require an additional EEG time-series recorded while the subject is performing a task inducing alpha desynchronisation. After computing the TF, the individual theta and alpha bands were defined as the frequency ranges [4, TF] Hz and [TF, 13] Hz, respectively.

### Connectivity measures

We considered weighted phase lag index (WPLI)^41^ and imaginary part of coherency (IMCOH)^42^ to assess synchronisation between pairs of EEG sensor recordings over time. In the following, we denote with *S* the number of EEG sensors and with *Z*_*i*_ (*f*) the Fourier transform at frequency *f* of the signal recorded by the *i*-th sensors. For each pair of signals *i, j* = 1, …, *S*, WPLI can be written ^9,41^ as

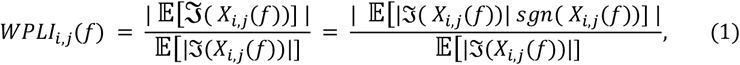

where 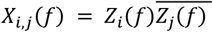 is the cross-spectrum between signals *i* and *j, ℑ* (*X*_*i,j*_(*f*)) is its imaginary part, and *sgn* is the sign function. WPLI assumes values from 0 to 1 representing lack or full connectivity, respectively.

The value of IMCOH between signal *i* and signal *j* at frequency *f* is defined as

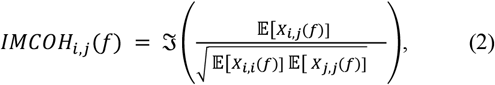

where again *X*_*i,j*_(*f*) is the cross-spectrum between signals *i* and *j*, and *ℑ*(*X*_*i,j*_(*f*)) is its imaginary part. Here we used the absolute values of IMCOH, which spans between 0 (lack of connectivity) and 1 (full connectivity). Hereafter, we will use |IMCOH| to refer to the absolute value of IMCOH.

The averaged value of *WPLI*_*i,j*_(*f*) and |*IMCOH*_*i,j*_(*f*)| over the individual theta and alpha bands were computed by exploiting routines from the MNE-Connectivity Python package (https://github.com/mne-tools/mne-connectivity). In detail, first, cross-spectrum densities estimation was performed using the multitaper method^43^ with digital prolate spheroidal sequence (DPSS)^44^ windows and then each connectivity measure was computed ad averaged across frequencies within a specific individual frequency band, returning one connectivity matrix of size *S × S* per individual and per frequency band. Further, in each connectivity map, the node degree was defined as the number of the edges connected to that node.

### EEG network statistical analysis

We used the Network Based Statistic (NBS) toolbox^45^ for statistical group-level analysis. Specifically, given two groups of individuals (either DLB and HC, or two subgroups of DLB patients with different core clinical features) and given an *S × S* connectivity matrix for each subject, NBS aims at identifying connected graphs that differentiate the two groups. Towards this end, first, a univariate one-tailed t-test is performed independently for each pair of sensors so as to test the null hypothesis that the mean connectivity between the two sensors is the same in the two groups against the alternative hypothesis of a greater mean connectivity in one of the groups. This procedure results in a *S × S* matrix of test statistic values that is thresholded by using the test statistic threshold *ts*_*th*_. Permutation testing is then performed to control the family-wise error rate. More in details, the suprathreshold connections are clustered in connected components *S*_*k*_ with *k* = 1, …, *K, K* being the number of identified components. The size *σ*_*k*_ of each *S*_*k*_ was computed by summing all the test statistic values corresponding to the connections belonging to *S*_*k*_. Then, subjects were randomly shuffled between the two groups and univariate statistical test, thresholding and connections-clustering were repeated for *L* times. For each permutation *l* = 1, …, *L*, the maximal subnetwork-statistic 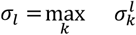 is stored and used to build an empirical null distribution of the maximal-subnetwork statistics. Eventually, the *p*-value of any observed connected component *S*_*k*_ of size *σ*_*k*_ is estimated as the percentage of permutations for which the maximal-subnetwork statistic is greater than *σ*_*k*_.

In our work, L was kept fixed for all tests, namely *L* = 5000, and the chosen statistical threshold was equal to *ts*_*th*_. Subnetworks were deemed significant if *p* < 0.05 for both WPLI and |IMCOH|. Statistically significant subnetworks were visualised by using the NetPlotBrain Python package^46^.

## Data Availability

All data produced in the present study are available upon reasonable request to the authors.

## Data availability

Data are available upon reasonable request to the corresponding author.

## Acknowledgments

S.S. acknowledges the support of the “Hub Life Science - Digital Health (LSH-DH) PNC-E3-2022-23683267 - Progetto DHEAL-COM - CUP: D33C22001980001”, granted by the Italian Ministero della Salute within the framework of the Piano Nazionale Complementare to the “PNRR Ecosistema Innovativo della Salute - Codice univoco investimento: PNC-E.3”. Mi.P. acknowledges the support of the European Union - NextGenerationEU and the Ministry of University and Research (MUR), National Recovery and Resilience Plan (NRRP), Mission 4, Component 2, Investment 1.5, project “RAISE - Robotics and AI for Socioeconomic Empowerment” (ECS00000035). Ma.P. acknowledges the support of the European Union - NextGenerationEU and the Ministry of University and Research (MUR), National Recovery and Resilience Plan (NRRP), project MNESYS (PE0000006) A Multiscale integrated approach to the study of the nervous system in health and disease (DN. 1553 11.10.2022). DA was supported by a grant from the Italian Ministry of health: Bando ricerca finalizzata RF-2021-12374240.

## Author information

Authors and Affiliations

Department of Mathematics (DIMA), University of Genoa, Genoa, Italy

Laura Carini, Sara Sommariva, Michele Piana

Department of Neuroscience, Rehabilitation, Ophthalmology, Genetics, Maternal and Child Health (DINOGMI), University of Genoa, Genoa, Italy

Dario Arnaldi, Francesco Famà, Laura Giorgetti, Pietro Mattioli, Beatrice Orso, Raffaele Mancini, Matteo Pardini

Neurophysiology Unit, IRCCS Ospedale Policlinico San Martino, Genoa, Italy

Dario Arnaldi, Francesco Famà, Pietro Mattioli

Clinical Neurology Unit, IRCCS Ospedale Policlinico San Martino, Genoa, Italy

Matteo Pardini

### Contributions

D.A. S.S. conceptualized and designed the study. L.C. and S.S. performed data analysis. S.S. D.A. Mi.P. provided input in the study plan and analyses. L.C. S.S. Mi.P. and D.A. wrote the first draft of the manuscript. D.A. F.F. L.G. P.M B.O. R.M. Ma.P collected clinical and instrumental data. All authors critically reviewed the manuscript.

Corresponding author: Sara Sommariva

## References

1. McKeith, I. G. et al. Diagnosis and management of dementia with Lewy bodies: Fourth consensus report of the DLB Consortium. Neurology 89, 88–100 (2017).

2. Joza, S. et al. Progression of clinical markers in prodromal Parkinson’s disease and dementia with Lewy bodies: a multicentre study. Brain 146, 3258–3272 (2023).

3. Arnaldi, D. et al. Presynaptic Dopaminergic Imaging Characterizes Patients with REM Sleep Behavior Disorder Due to Synucleinopathy. Ann. Neurol. 95, 1178–1192 (2024).

4. Simuni, T. et al. A biological definition of neuronal α-synuclein disease: towards an integrated staging system for research. Lancet Neurol. 23, 178–190 (2024).

5. Höglinger, G. U. et al. A biological classification of Parkinson’s disease: the SynNeurGe research diagnostic criteria. Lancet Neurol. 23, 191–204 (2024).

6. Kucikova, L. et al. A systematic literature review of fMRI and EEG resting-state functional connectivity in Dementia with Lewy Bodies: Underlying mechanisms, clinical manifestation, and methodological considerations. Ageing Res. Rev. 93, 102159 (2024).

7. Schumacher, J. et al. Functional connectivity in dementia with Lewy bodies: A within- and between-network analysis. Hum. Brain Mapp. 39, 1118–1129 (2018).

8. Wada, A. et al. Differentiating Alzheimer’s Disease from Dementia with Lewy Bodies Using a Deep Learning Technique Based on Structural Brain Connectivity. Magn. Reson. Med. Sci. 18, 219–224 (2019).

9. Mehraram, R. et al. Functional and structural brain network correlates of visual hallucinations in Lewy body dementia. Brain 145, 2190–2205 (2022).

10. Vallesi, A. et al. Resting-state EEG spectral and fractal features in dementia with Lewy bodies with and without visual hallucinations. Clin. Neurophysiol. 168, 43–51 (2024).

11. Roascio, M. et al. Phase and amplitude electroencephalography correlations change with disease progression in people with idiopathic rapid eye-movement sleep behavior disorder. Sleep 45, (2022).

12. Jeong, E. et al. EEG-based machine learning models for the prediction of phenoconversion time and subtype in isolated rapid eye movement sleep behavior disorder. Sleep 47, (2024).

13. Klimesch, W., Schimke, H. & Pfurtscheller, G. Alpha frequency, cognitive load and memory performance. Brain Topogr. 5, 241–251 (1993).

14. Nunez, P. L., Reid, L. & Bickford, R. G. The relationship of head size to alpha frequency with implications to a brain wave model. Electroencephalogr. Clin. Neurophysiol. 44, 344–352 (1978).

15. Stylianou, M. et al. Quantitative electroencephalography as a marker of cognitive fluctuations in dementia with Lewy bodies and an aid to differential diagnosis. Clin. Neurophysiol. 129, 1209–1220 (2018).

16. Klimesch, W. EEG alpha and theta oscillations reflect cognitive and memory performance: a review and analysis. Brain Res. Rev. 29, 169–195 (1999).

17. Moretti, D. Individual analysis of EEG frequency and band power in mild Alzheimer’s disease. Clin. Neurophysiol. 115, 299–308 (2004).

18. Vallarino, E. et al. Transfreq: A Python package for computing the theta-to-alpha transition frequency from resting state electroencephalographic data. Hum. Brain Mapp. 43, 5095–5110 (2022).

19. Lex, A., Gehlenborg, N., Strobelt, H., Vuillemot, R. & Pfister, H. Upset: visualization of intersecting sets. IEEE Trans. Vis. Comput. Graph. 20, 1983–1992 (2014).

20. Habich, A., Wahlund, L.-O., Westman, E., Dierks, T. & Ferreira, D. (Dis-)Connected Dots in Dementia with Lewy Bodies-A Systematic Review of Connectivity Studies. Mov. Disord. 38, 4–15 (2023).

21. Babiloni, C. et al. Abnormalities of functional cortical source connectivity of resting-state electroencephalographic alpha rhythms are similar in patients with mild cognitive impairment due to Alzheimer’s and Lewy body diseases. Neurobiol. Aging 77, 112–127 (2019).

22. Bonanni, L. et al. EEG Markers of Dementia with Lewy Bodies: A Multicenter Cohort Study. J Alzheimers Dis 54, 1649–1657 (2016).

23. Lachaux, J. P., Rodriguez, E., Martinerie, J. & Varela, F. J. Measuring phase synchrony in brain signals. Hum. Brain Mapp. 8, 194–208 (1999).

24. Fries, P. A mechanism for cognitive dynamics: neuronal communication through neuronal coherence. Trends Cogn Sci (Regul Ed) 9, 474–480 (2005).

25. Chialvo, D. R. Emergent complex neural dynamics. Nat. Phys. 6, 744–750 (2010).

26. Bassett, D. S. & Bullmore, E. T. Small-World Brain Networks Revisited. Neuroscientist 23, 499–516 (2017).

27. Leodori, G. et al. Effective connectivity abnormalities in Lewy body disease with visual hallucinations. Clin. Neurophysiol. 156, 156–165 (2023).

28. Pusil, S. et al. Hypersynchronization in mild cognitive impairment: the “X” model. Brain 142, 3936–3950 (2019).

29. Frantzidis, C. A. et al. Functional disorganization of small-world brain networks in mild Alzheimer’s Disease and amnestic Mild Cognitive Impairment: an EEG study using Relative Wavelet Entropy (RWE). Front. Aging Neurosci. 6, 224 (2014).

30. Schutte, M. J. L. et al. Functional connectome differences in individuals with hallucinations across the psychosis continuum. Sci. Rep. 11, 1108 (2021).

31. Kenny, E. R., O’Brien, J. T., Firbank, M. J. & Blamire, A. M. Subcortical connectivity in dementia with Lewy bodies and Alzheimer’s disease. Br. J. Psychiatry 203, 209–214 (2013).

32. Kenny, E. R., Blamire, A. M., Firbank, M. J. & O’Brien, J. T. Functional connectivity in cortical regions in dementia with Lewy bodies and Alzheimer’s disease. Brain 135, 569–581 (2012).

33. Schumacher, J. et al. Functional connectivity of the nucleus basalis of Meynert in Lewy body dementia and Alzheimer’s disease. Int. Psychogeriatr. 33, 89–94 (2021).

34. Babiloni, C. et al. Abnormalities of resting-state functional cortical connectivity in patients with dementia due to Alzheimer’s and Lewy body diseases: an EEG study. Neurobiol. Aging 65, 18–40 (2018).

35. Morbelli, S. et al. Metabolic patterns across core features in dementia with lewy bodies. Ann. Neurol. 85, 715–725 (2019).

36. Orso, B. et al. Progression trajectories from prodromal to overt synucleinopathies: a longitudinal, multicentric brain [18F]FDG-PET study. npj Parkinsons Disease 10, 200 (2024).

37. Mattioli, P. et al. Cuneus/precuneus as a central hub for brain functional connectivity of mild cognitive impairment in idiopathic REM sleep behavior patients. Eur. J. Nucl. Med. Mol. Imaging 48, 2834–2845 (2021).

38. Gramfort, A. et al. MEG and EEG data analysis with MNE-Python. Front. Neurosci. 7, 267 (2013).

39. Offner, F. F. The EEG as potential mapping: the value of the average monopolar reference. Electroencephalogr. Clin. Neurophysiol. 2, 213–214 (1950).

40. Jutten, C. & Herault, J. Blind separation of sources, part I: An adaptive algorithm based on neuromimetic architecture. Signal Processing 24, 1–10 (1991).

41. Vinck, M., Oostenveld, R., van Wingerden, M., Battaglia, F. & Pennartz, C. M. A. An improved index of phase-synchronization for electrophysiological data in the presence of volume-conduction, noise and sample-size bias. Neuroimage 55, 1548–1565 (2011).

42. Nolte, G. et al. Identifying true brain interaction from EEG data using the imaginary part of coherency. Clin. Neurophysiol. 115, 2292–2307 (2004).

43. Thomson, D. J. Spectrum estimation and harmonic analysis. Proc. IEEE 70, 1055–1096 (1982).

44. Slepian, D. Prolate Spheroidal Wave Functions, Fourier Analysis, and Uncertainty-V: The Discrete Case. Bell System Technical Journal 57, 1371–1430 (1978).

45. Zalesky, A., Fornito, A. & Bullmore, E. T. Network-based statistic: identifying differences in brain networks. Neuroimage 53, 1197–1207 (2010).

46. Fanton, S. & Thompson, W. H. NetPlotBrain: A Python package for visualizing networks and brains. Netw. Neurosci. 7, 461–477 (2023).

