## Supplementary material for "Network based statistics shows that rem sleep behavior disorder and visual hallucinations increase functional connectivity in early dementia with Lewy bodies": Figure 1 and 2

### Supplementary materials

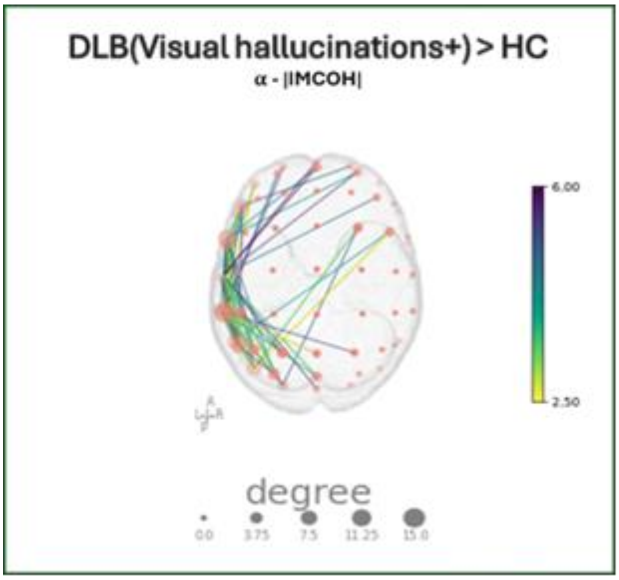


Supplementary Figure 1. Statistically significant differential network provided by NBS when testing for an increased value of |IMCOH| within the individual alpha-band from DLB patients with visual hallucinations (DLB(Visual hallucinations+)) than in HC. As in Figure 3, edge colours represent the t-values provided by the first step of NBS while each node size is proportional to its degree

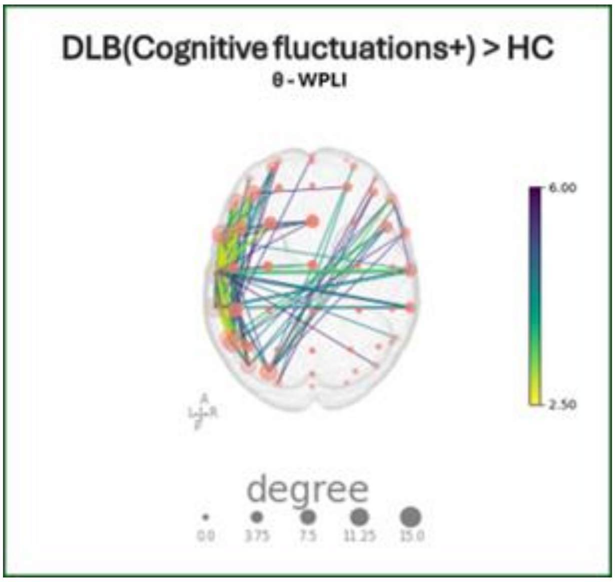


Supplementary Figure 2. Statistically significant differential network provided by NBS when testing for an increased value of WPLI within the individual theta-band from DLB patients with cognitive fluctuations (DLB(Cognitive fluctuations+)) than in HC. As in Figure 3, edge colours represent the t-values provided by the first step of NBS while each node size is proportional to its degree
